# Association of Body Composition With Tumor Proteomics and Survival in Patients With Clear Cell Renal Cell Carcinoma

**DOI:** 10.1101/2025.09.19.25336108

**Authors:** Cuthbert Mario Mahenge, Rand Talal Akasheh, Xuan Nguyen, Ting-Yuan David Cheng

## Abstract

**Background:** Prognoses for patients with clear cell renal cell carcinoma (ccRCC) are associated with complex interactions between tumor and patient characteristics. This study investigated associations between body composition and tumor proteomics and their interaction with survival among patients with ccRCC.

**Methods:** Data from 178 patients in the TCGA-KIRC project were analyzed to assess adipose and skeletal muscle tissue areas at the third lumbar vertebra of diagnostic computed tomography scan images. Patients were classified into four body composition types: high muscle with low adiposity; high muscle with high adiposity; low muscle with low adiposity; and low muscle with high adiposity. Proteins with differential expression were screened for interactions with body composition type on survival. Linear regression was used to assess associations, and Cox regression models—adjusted for age, tumor stage, sex, race, and ethnicity—were utilized for survival analysis.

**Results:** Patients having low muscle with low adiposity exhibited worse survival than those having high muscle with high adiposity (hazard ratio, 3.74 [95% CI, 1.69–8.27]). Low muscle with low adiposity was also associated with increased expression of P-cadherin and decreased expression of DIRAS3 (P<0.05; false discovery rate–corrected P<0.1), both associated with poor survival in the entire KIRC cohort. Among patients having low muscle with high adiposity, high (vs. low) PREX1 expression was associated with 15.8-fold (95% CI, 3.08–80.78) increased mortality.

**Conclusion:** Body composition is associated with differential expression of proteins and survival in ccRCC.

**Impact:** Body composition and tumor proteomics may be prognostic biomarkers and therapeutic targets in ccRCC.

## Introduction

Clear cell renal cell carcinoma (ccRCC) is the most prevalent type of kidney cancer, accounting for about 80% of all kidney cancer cases in the United States (1). The incidence of ccRCC has been steadily rising, with an estimated 81,000 new diagnoses and nearly 15,000 deaths in 2023 (1). This malignant tumor is more common in men, with a male:female incidence ratio of approximately 2:1 (2,3). The risk increases with age, peaking in individuals between 60 and 70 years of age (3). As the global population ages, the age-related susceptibility to ccRCC makes it increasingly critical to conduct comprehensive research to improve prevention strategies and patient outcomes. Currently, there are no effective screening programs or validated tumor markers for the early detection of ccRCC (4,5), in part due to the heterogeneity of the cancer (6). This leads to the majority of diagnoses occurring at advanced stages, for which the 5-year survival rate is less than 18% despite recent healthcare advances (7). Poor survival can also be attributed to the strong resistance to conventional chemotherapy and radiotherapy of these tumors, presenting profound challenges and complicating treatment options that can be addressed by targeted therapy (4). The prognosis for patients with ccRCC mainly relies on histological and clinical data (8).

Nonetheless, there is a growing consensus in the health community on the need to enhance patient stratification methods by incorporating characteristics that influence both risk and survival outcomes.

Obesity has been well established as a primary risk factor for kidney cancers (9), likely due to chronic inflammation and alterations in the insulin signaling pathway (10).

Despite the increased risk, obesity is associated with less aggressive pathologic features and a more favorable prognosis among patients with ccRCC (11–13). Most studies assessing this association used body mass index (BMI) as a measure of adiposity. However, BMI does not accurately estimate the quantities of adipose and muscle, important body composition components that may play important yet different roles in patient outcomes. Some studies have investigated the associations between body fat and muscle mass with ccRCC outcomes (14–16). Specifically, low muscle mass and low subcutaneous adipose tissue have been associated with shorter overall survival (16–18). However, findings examining visceral adipose tissue have been inconsistent, with some studies associating lower quantities with shorter survival, while others associate higher quantities and radiodensity to poor outcomes (17,18). These findings underscore the complex relationships between body composition components and ccRCC, highlighting the need for further research. Given that most patients with ccRCC undergo computed tomography (CT) scans for diagnosis, cancer staging, and follow-up, body composition assessment can be readily performed using available images. By analyzing CT scans at the third lumbar (L3) vertebral level, muscle and adipose tissue areas, which strongly correlate with the total quantities of these tissues in the whole body, can be accurately measured (19).

The molecular profile of ccRCC also significantly affects patient prognosis (20). Historically, ccRCC was primarily associated with mutations in the VHL gene, but recent studies have shown that additional genetic alterations and molecular signatures correlate with prognosis, metastasis, and response to therapy (21–23). Thus, there is a pressing need to assess the combined effects of tumor signaling pathways and body composition profiles in patients with ccRCC.

This study aims to investigate the role of body composition components and tumor protein expression in the survival of patients with ccRCC. Findings from this research would advance the understanding of the impact of body composition on ccRCC progression and integrate body composition and tumor protein markers in risk stratification.

## Materials and Methods

### Study Participants

Data for patients were obtained from The Cancer Genome Atlas - Kidney Renal Clear Cell Carcinoma (TCGA-KIRC) cohort. TCGA is a collaborative initiative spearheaded by the National Cancer Institute and the National Human Genome Research Institute. This project combines research data from multiple institutions to map and understand the germline and somatic alterations in cancers. The clinical, pathologic, and molecular data are stored at the Genomic Data Commons data portal, and the imaging data are stored at the Cancer Imaging Archive (24). We obtained the CT scans in the archive from routine clinical imaging procedures conducted immediately before the pathologic diagnosis or a follow-up image taken within a 3-month window after the initial diagnosis.

The TCGA-KIRC has 537 patients with clinical data. Among them, 300 patients were excluded from the present study for the lack of CT scan images, and 38 patients were excluded for having ineligible CT scans due to a part of the body being cropped out of the image, the image having poor resolution, or the axial (transverse) CT scan lacking an L3 slice image. The remaining 199 patients with CT scan images were included in the survival analysis. Among these patients, 21 with missing proteomic data were excluded. Therefore, 178 patients were included in the analysis of the association between body composition and protein expression. In cases of multiple CT scans being available for a patient, the earliest eligible scan was used. Proteins in the TCGA-KIRC data were assayed by reverse-phase protein array. For the analysis, 233 proteins with non-missing values for KIRC patients present in the proteomic dataset were included. **Supplemental Figure S1** shows the flowchart of patient selection.

### CT image analysis

An image corresponding to the L3 vertebra for each patient was selected using Sante DICOM Viewer Lite Version 3.1.5 (64-bit) (Santesoft LTD; Nicosia, Cyprus). Images were analyzed with SliceOmatic, version 5.0 revision 11b (Tomovision; Montreal, Canada) to quantify visceral adipose tissue (VAT), subcutaneous adipose tissue (SAT), intramuscular adipose tissue (IMAT), and total skeletal muscle (TSM) areas. Tissue identification relied on Hounsfield unit values and anatomical positioning (**Supplemental Table S1**) (25). A trained researcher performed all annotations, which a board-certified radiologist subsequently reviewed for confirmation. Total adipose tissue (TAT) was calculated by summing VAT, SAT, and IMAT values. The Contal and O’Quigley method was used to determine the optimal cutoff values for TAT and TSM quantities that best separate patients into two distinct risk groups based on overall survival. This analysis was performed using the *surv_cutpoint* function in the *survminer* package (version 0.4.9) in R (version 4.4.0; R Core Team, 2024). The optimal cutoff values for TAT and TSM area quantities were calculated separately for male and female patients because of significant differences in adipose and muscle mass by sex (26,27). Patients were then classified into four mutually exclusive body composition groups based on the areas of the TSM and TAT categories: (1) high muscle with high adiposity, (2) high muscle with low adiposity, (3) low muscle with high adiposity, and (4) low muscle with low adiposity.

### Statistical analysis

Descriptive statistics were computed to summarize demographic and clinical data. Means and standard deviations were used for continuous variables, whereas frequencies and percentages were used for categorical variables. Survival analysis was conducted using Kaplan-Meier curves and adjusted Cox proportional hazards models. The group with the best overall survival was selected as the reference group for estimating hazard ratios (HRs) and their corresponding 95% confidence intervals (CIs). The analysis was adjusted for age, sex, race, ethnicity, and cancer stage.

To assess the association between the body composition types and tumor protein expression (in log_2_ normalized values), linear regression analysis adjusted for age, sex, race, ethnicity, and cancer stage was employed, with the high muscle with high adiposity type as the reference group. False discovery rate (FDR) adjustment was performed using the Benjamini-Hochberg method. The FDR cutoff was set at 0.2. We investigated the association of proteins showing differential expression (nominal p<0.05) with survival in combination with the body composition types. Optimal cutoff values for protein expression levels were determined using the Contal and O’Quigley method to effectively stratify patients into two distinct risk groups (high vs. low) based on overall survival. This analysis was performed using the *surv_cutpoint* function in the *survminer* package in R software. The corresponding survival curves were then generated using the *ggsurvplot* function in the same package. Cox proportional hazards models were constructed, incorporating an interaction term between body composition groups and proteins.

All statistical analyses were performed using SAS 9.4 (SAS Institute Inc; North Carolina, USA) and R (version 4.4.0; R Core Team, 2024). The volcano plots were generated using GraphPad Prism version 10.1.2 (GraphPad Software; MA, USA).

## Data Availability

The CT images analyzed in this study were obtained from the Cancer Imaging Archive at TCIA: <u>TCGA-KIRC</u>(24); Clinical and protein data are available in the National Cancer Institute’s Genomic Data Commons at <u>GDC: TCGA-KIRC</u>(28) and at <u>MD Anderson Cancer Center’s The Cancer Proteome Atlas</u>, respectively. Data generated by the authors is available upon request.

## Results

### Patient characteristics

**Table 1** summarizes the demographic and clinical characteristics of the patients with clinical, proteomic, and CT scan data based on their body composition distribution. Their age at diagnosis ranged from 26 to 88 years, with a mean of 59.6 years. Most patients (94.4%) were White, with non-Hispanic or Latino being the predominant ethnicity (94.3% among those with a non-missing value). Male patients comprised approximately two-thirds (66.9%) of the sample population. Most patients (51.7%) were diagnosed as having stage I ccRCC, with 8.4%, 25.3%, and 14.6% of patients having stages II, III, and IV, respectively. The majority of patients (57.9%) were categorized as being in the high muscle with high adiposity body composition type, with 9.0% in the high muscle with low adiposity type, 20.8% in the low muscle with low adiposity type, and 12.4% in the low muscle with high adiposity type.

**Table 1.** Characteristics of Patients Included in Analyses.

| Characteristic | Total | Low muscle with high adiposity | Low muscle with low adiposity | High muscle with high adiposity | High muscle with low adiposity | P-value |
| --- | --- | --- | --- | --- | --- | --- |
| **No. (%) in group** | 178 (100%) | 22 (12.4%) | 37 (20.8%) | 103 (57.9%) | 16 (9.0%) |  |
| **Age** |  |  |  |  |  | <.001<sup>1</sup> |
| Mean (SD), y | 59.6 (13.2) | 68.4 (10.6) | 65.7 (13.2) | 56.3 (12.5) | 55.2 (10.5) |  |
| **Sex, n (%)** |  |  |  |  |  | <.001<sup>2</sup> |
| Female | 59 (33.1%) | 11 (50.0%) | 29 (78.4%) | 11 (10.7%) | 8 (50.0%) |  |
| Male | 119 (66.9%) | 11 (50.0%) | 8 (21.6%) | 92 (89.3%) | 8 (50.0%) |  |
| **Race, n (%)** |  |  |  |  |  | 0.116<sup>2</sup> |
| Asian | 2 (1.1%) | 0 (0%) | 1 (2.7%) | 0 (0.0%) | 1 (6.3%) |  |
| Black or African American | 8 (4.5%) | 0 (0%) | 0 (0%) | 7 (6.8%) | 1 (6.3%) |  |
| White | 168 (94.4%) | 22 (100%) | 36 (97.3%) | 96 (93.2%) | 14 (87.5%) |  |
| **Ethnicity, n (%)** |  |  |  |  |  | 0.718<sup>2</sup> |
| Hispanic or Latino | 7 (3.9%) | 1 (4.5%) | 1 (2.7%) | 4 (3.9%) | 1 (6.3%) |  |
| Not Hispanic or Latino | 116 (65.2%) | 16 (72.7%) | 26 (70.3%) | 62 (60.2%) | 12 (75.0%) |  |
| Not reported | 55 (30.9%) | 5 (22.7%) | 10 (27.0%) | 37 (35.9%) | 3 (18.8%) |  |
| **AJCC pathologic stage, n (%)** |  |  |  |  |  | <.001<sup>2</sup> |
| I | 92 (51.7%) | 11 (50.0%) | 11 (29.7%) | 63 (61.2%) | 7 (43.8%) |  |
| II | 15 (8.4%) | 1 (4.5%) | 0 (0%) | 14 (13.6%) | 0 (0%) |  |
| III | 45 (25.3%) | 5 (22.7%) | 15 (40.5%) | 18 (17.5%) | 7 (43.8%) |  |
| IV | 26 (14.6%) | 5 (22.7%) | 11 (29.7%) | 8 (7.8%) | 2 (12.5%) |  |
| **Vital status, n (%)** |  |  |  |  |  | <.001<sup>2</sup> |
| Alive | 122 (68.5%) | 15 (68.2%) | 13 (35.1%) | 84 (81.6%) | 10 (62.5%) |  |
| Dead | 56 (31.5%) | 7 (31.8%) | 24 (64.9%) | 19 (18.4%) | 6 (37.5%) |  |
| **Visceral adipose tissue area** |  |  |  |  |  | <.001<sup>1</sup> |
| Mean (SD), cm<sup>2</sup> | 191.6 (117.93) | 243.6 (133.32) | 77.9 (44.38) | 240.5 (97.10) | 67.4 (40.57) |  |
| **Intramuscular adipose tissue area** |  |  |  |  |  | <.001<sup>1</sup> |
| Mean (SD), cm<sup>2</sup> | 9.2 (6.42) | 12.6 (7.60) | 9.3 (6.95) | 9.2 (5.81) | 4.0 (3.66) |  |
| **Subcutaneous adipose tissue area** |  |  |  |  |  | <.001<sup>1</sup> |
| Mean (SD), cm<sup>2</sup> | 234.0 (119.30) | 297.8 (142.44) | 161.3 (56.17) | 261.7 (117.61) | 136.7 (59.50) |  |
| **Total adipose tissue area** |  |  |  |  |  | <.001<sup>1</sup> |
| Mean (SD), cm<sup>2</sup> | 434.8 (198.76) | 554.0 (189.00) | 248.5 (84.46) | 511.4 (168.22) | 208.1 (70.82) |  |
| **Muscle area** |  |  |  |  |  | <.001<sup>1</sup> |
| Mean (SD), cm<sup>2</sup> | 159.9 (41.74) | 125.6 (19.46) | 108.4 (18.87) | 186.4 (27.94) | 155.5 (27.35) |  |

<sup>1</sup>Kruskal-Wallis P-value; <sup>2</sup>Chi-Square P-value; Abbreviation: AJCC, American Joint Committee on Cancer

### Body composition and survival

The identified cutoff values for TSM were 153.00 cm^2^ for male patients and 120.70 cm^2^ for female patients (**Supplemental Figure S2**). The cutoff values for TAT were 239.11 cm^2^ for male patients and 383.05 cm^2^ for female patients (**Supplemental Figure S3**). Survival analysis of the TAT groups presented in **Table 2** showed that individuals in the low TAT group had more than three times the mortality risk of those in the high TAT group (HR, 3.24 [95% CI, 1.56–6.74; P = 0.002) (**Figure 1A**). Total muscle groups did not demonstrate a significant association with overall survival (low vs. high; HR, 1.15 [95% CI, 0.58–2.30]; P = 0.685) (**Table 2** and **Figure 1B**). Compared with the high muscle with high adiposity body composition type, which had the best overall survival, the low muscle with low adiposity type experienced the worst overall survival outcomes (HR, 3.74 [95% CI, 1.69–8.27]; P = 0.001), followed by the high muscle with low adiposity type (HR, 3.20; [95% CI, 1.22–8.42]; P = 0.018) (**Table 2** and **Figure 1C**). The associations of other covariates with mortality is presented in **Supplemental Table S2**. **Body composition and tumor protein expression**

**Figure 1:**
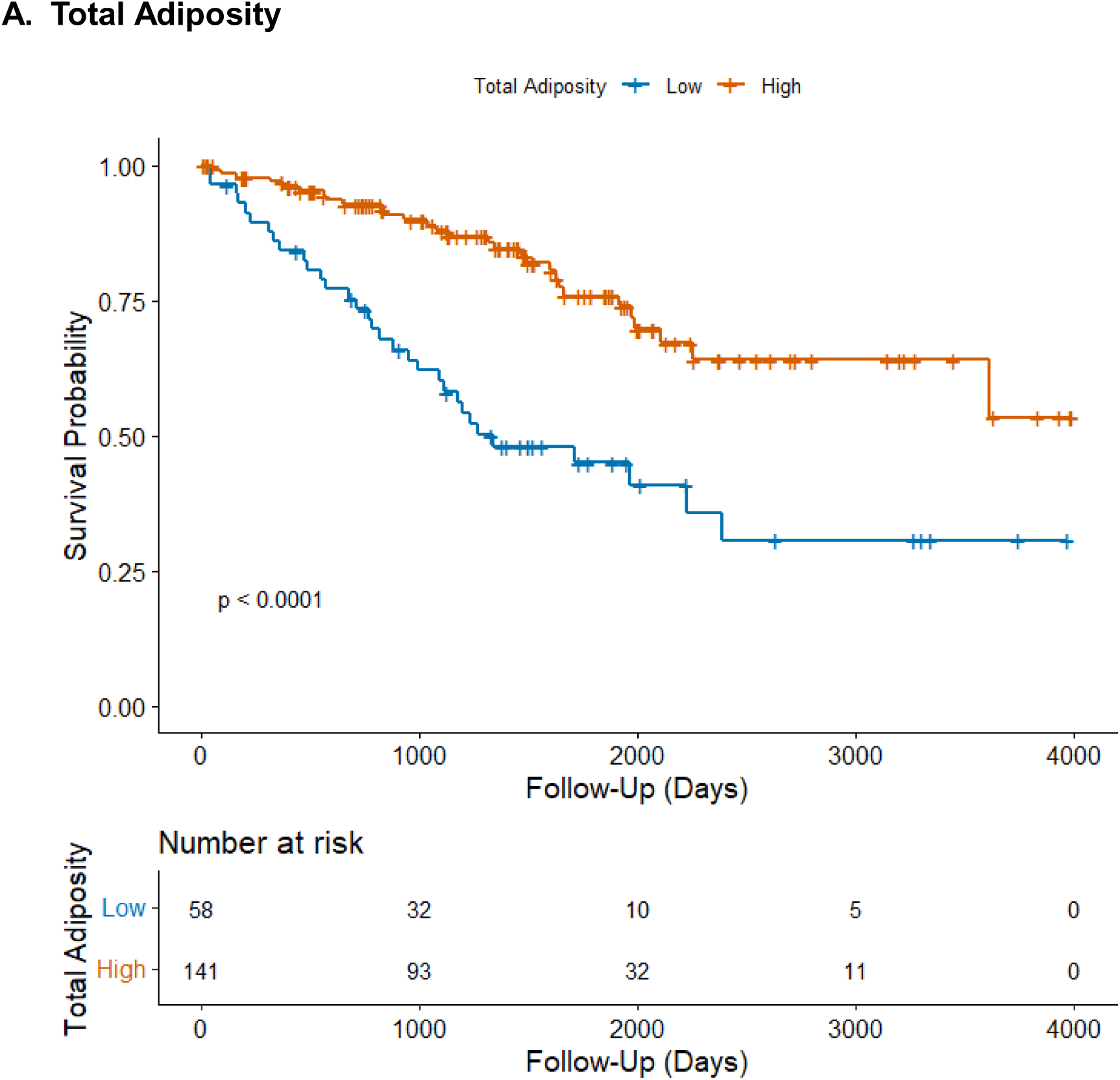
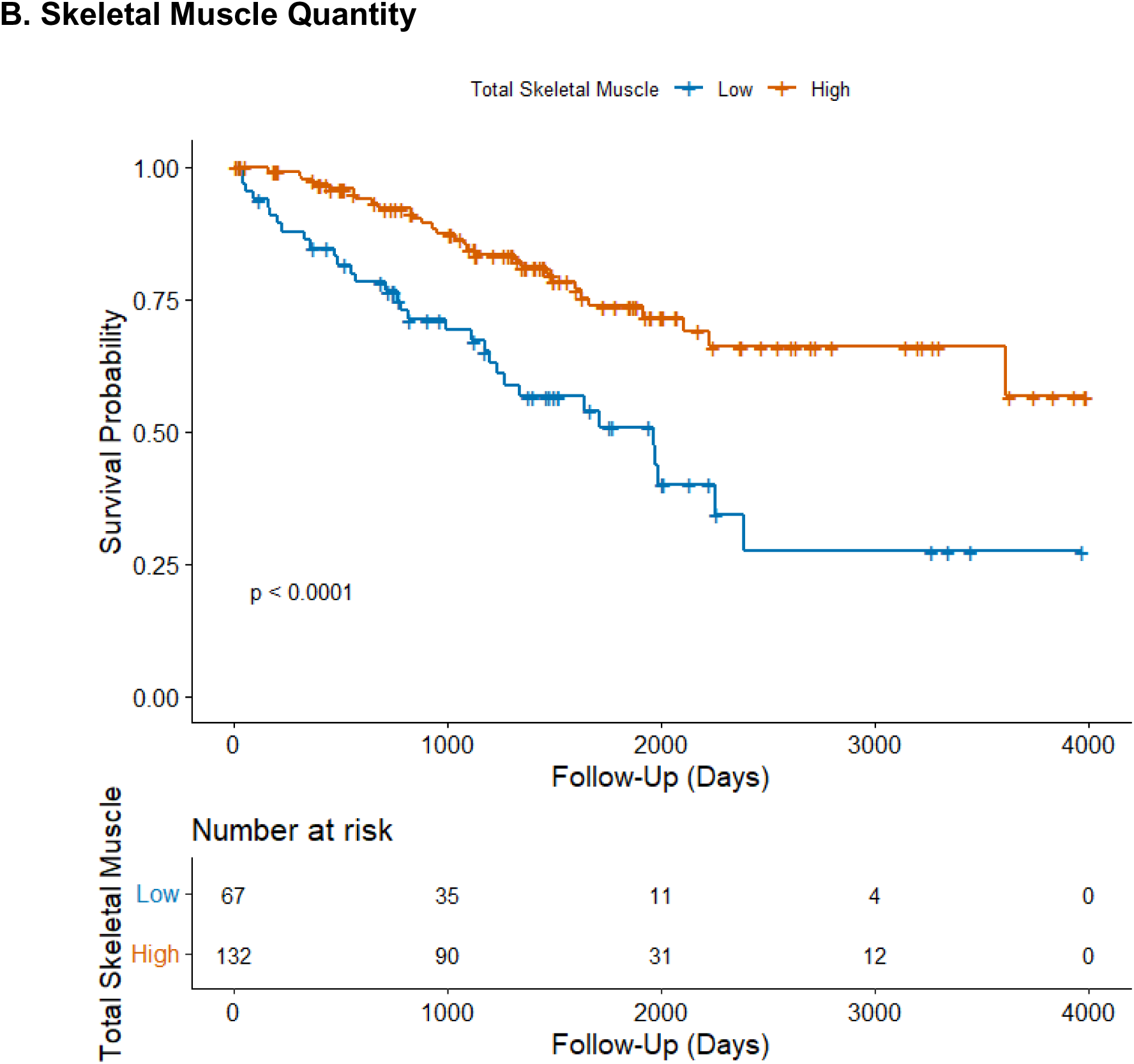
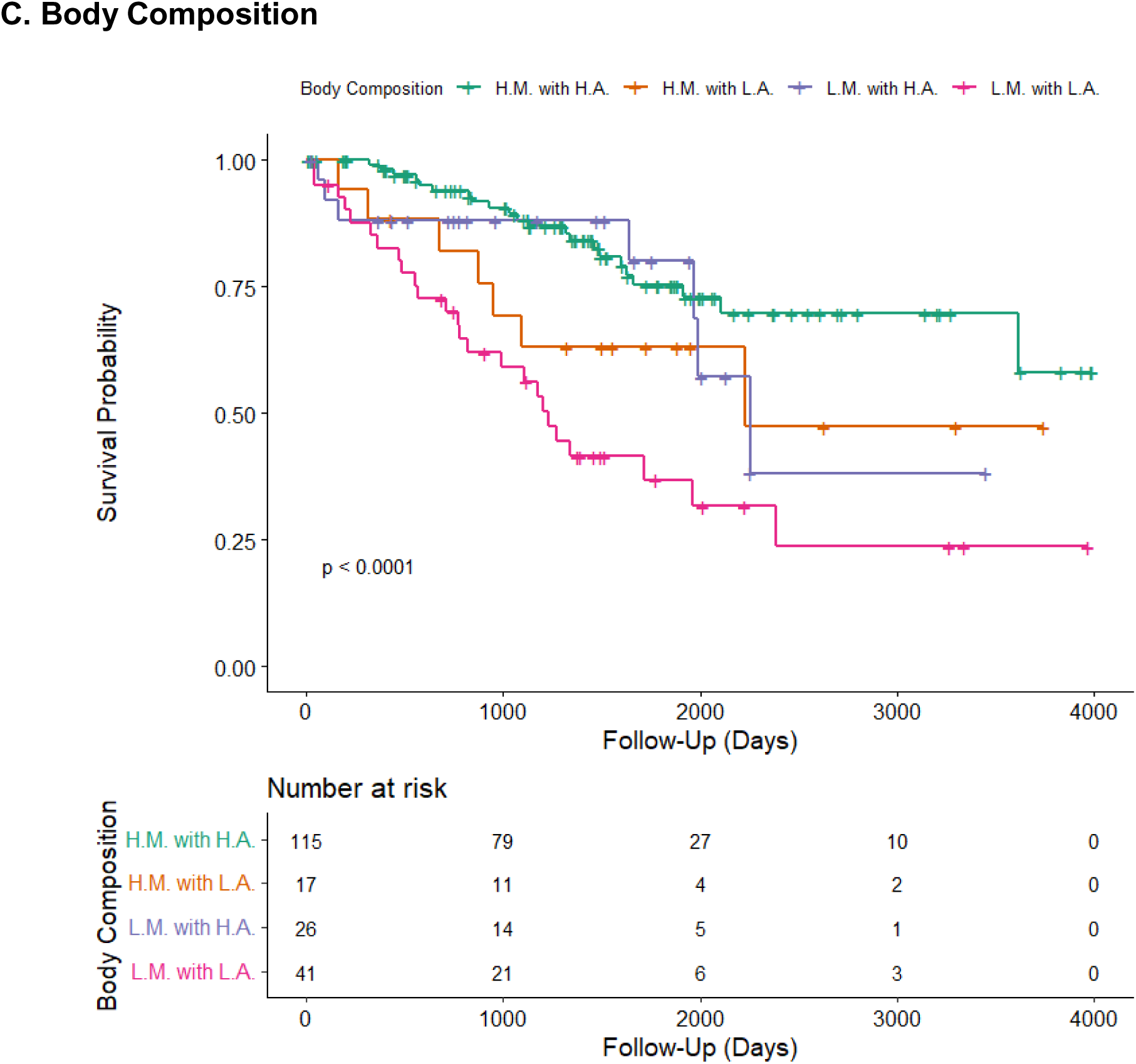
**Kaplan-Meier Curves Exploring Survival Among Participants Based on Body Composition Component and Category.**

**Table 2.** Mortality Risk by Body Composition.

|  | Events/Total | Hazard Ratio<br>(95% CI) | P-value |
| --- | --- | --- | --- |
| **Total Adiposity\*** |  |  |  |
| Low | 32/58 | 3.24 (1.56–6.74) | 0.002<sup>1</sup> |
| High | 29/141 | Reference |  |
| **Skeletal Muscle\*\*** |  |  |  |
| Low | 32/67 | 1.15 (0.58–2.30) | 0.685<sup>1</sup> |
| High | 29/132 | Reference |  |
| **Body Composition\*\*\*** |  |  | 0.005<sup>2</sup> |
| High muscle with low adiposity | 7/17 | 3.20 (1.22–8.42) | 0.018<sup>1</sup> |
| Low muscle with low adiposity | 25/41 | 3.74 (1.69–8.27) | 0.001<sup>1</sup> |
| Low muscle with high adiposity | 7/26 | 1.14 (0.45–2.91) | 0.786<sup>1</sup> |
| High muscle with high adiposity | 22/115 | Reference |  |
| <sup>1</sup>Covariate Wald P-value; <sup>2</sup>Type 3 Wald P-value<br>*Total adiposity: Adjusted for stage, sex, race, ethnicity, age, and Total Muscle area.<br>**Skeletal muscle: Adjusted for stage, sex, race, ethnicity, age, and Total Adiposity.<br>***Body composition: Adjusted for stage, sex, race, ethnicity, and age<br>Abbreviation: CI, confidence interval |  |  |  |

High (vs. low) TSM levels were associated with higher expression of androgen receptor (log_2_ fold change, 0.2; P = 0.02), carbonic anhydrase IX (CA9) (log_2_ fold change, 0.3; P = 0.037), interferon regulatory factor 1 (IRF-1) (log_2_ fold change, 0.1; P = 0.034), and BCL2-related protein A1 (BCL2A1) (log_2_ fold change, 0.2; P = 0.038), but lower expression of P-cadherin (log_2_ fold change, −0.2; P = 0.025), caspase-3 (log_2_ fold change, −0.1; P = 0.018), and phosphorylated myosin IIa at serine 1943 (phospho-myosin IIa (S1943) (log_2_ fold change, −0.2; P = 0.024) (**Figure 2A**).

**Figure 2.**
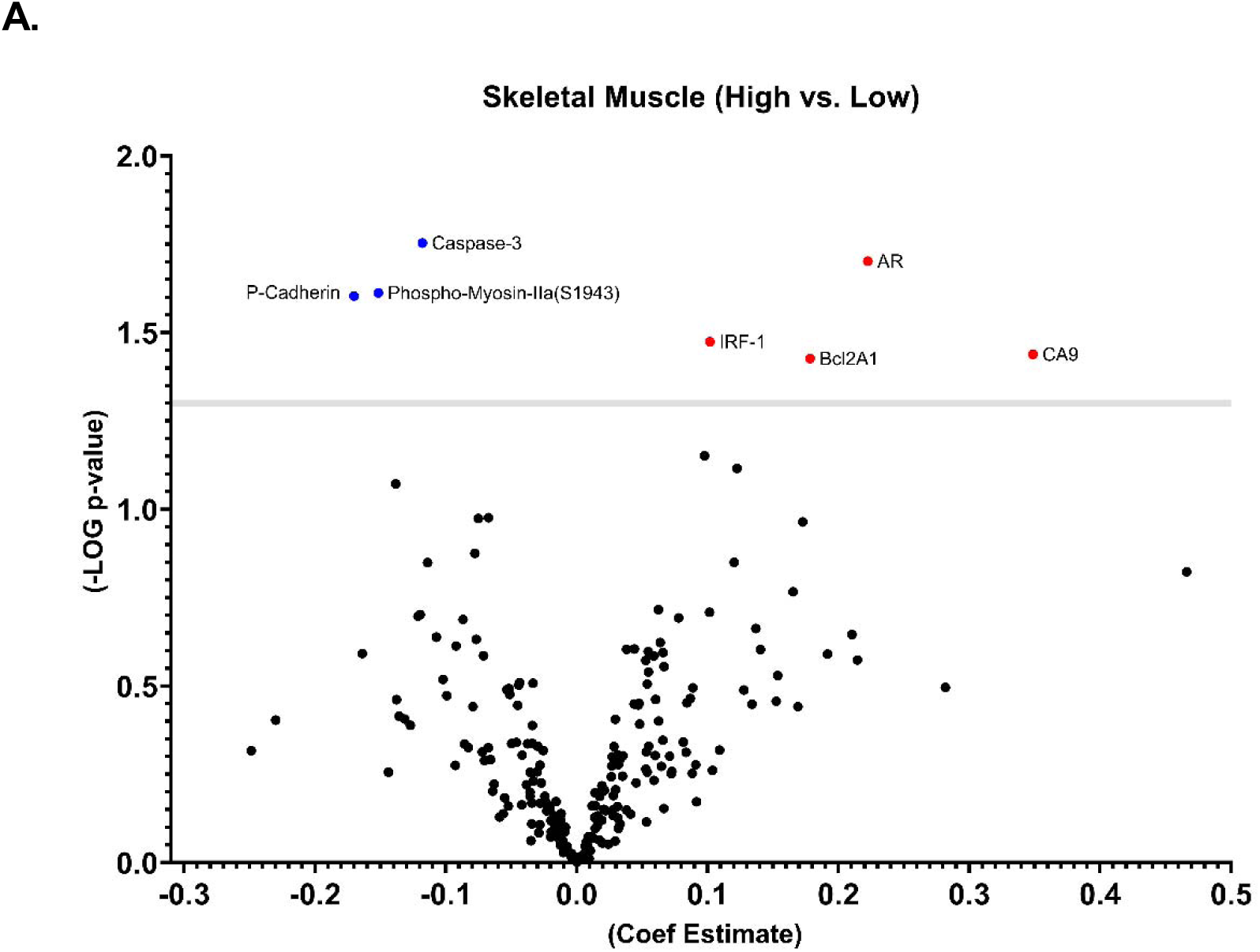
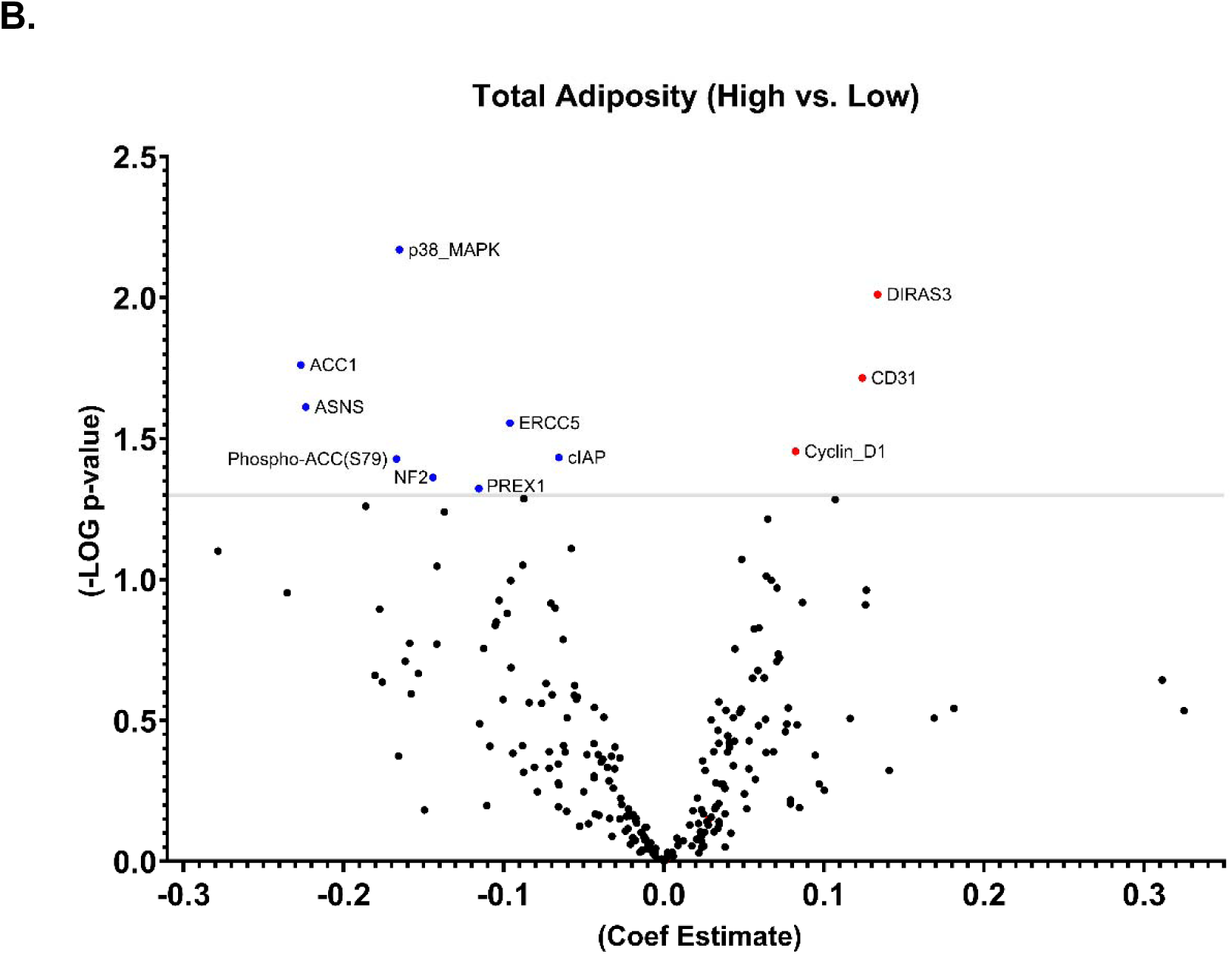
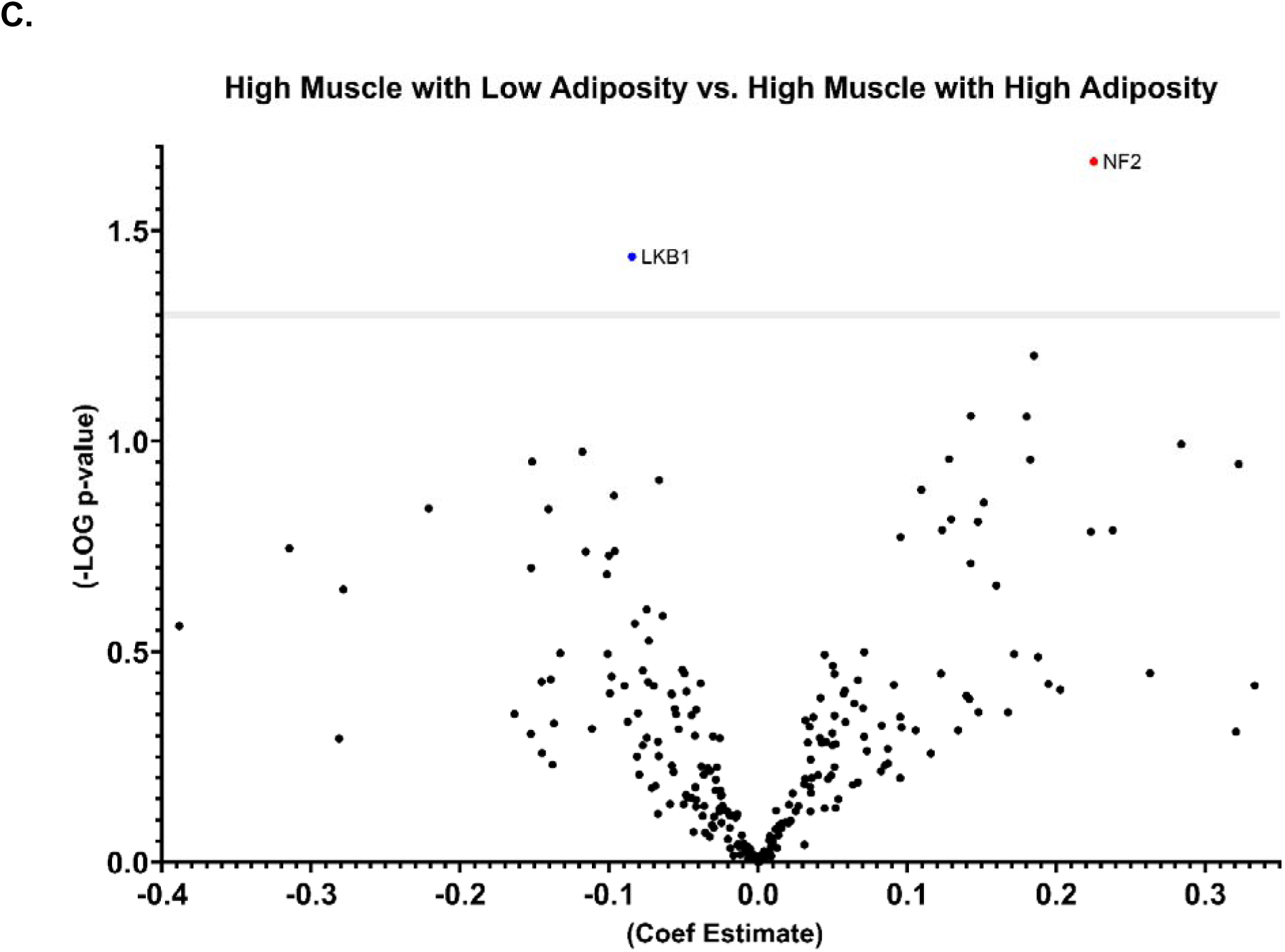
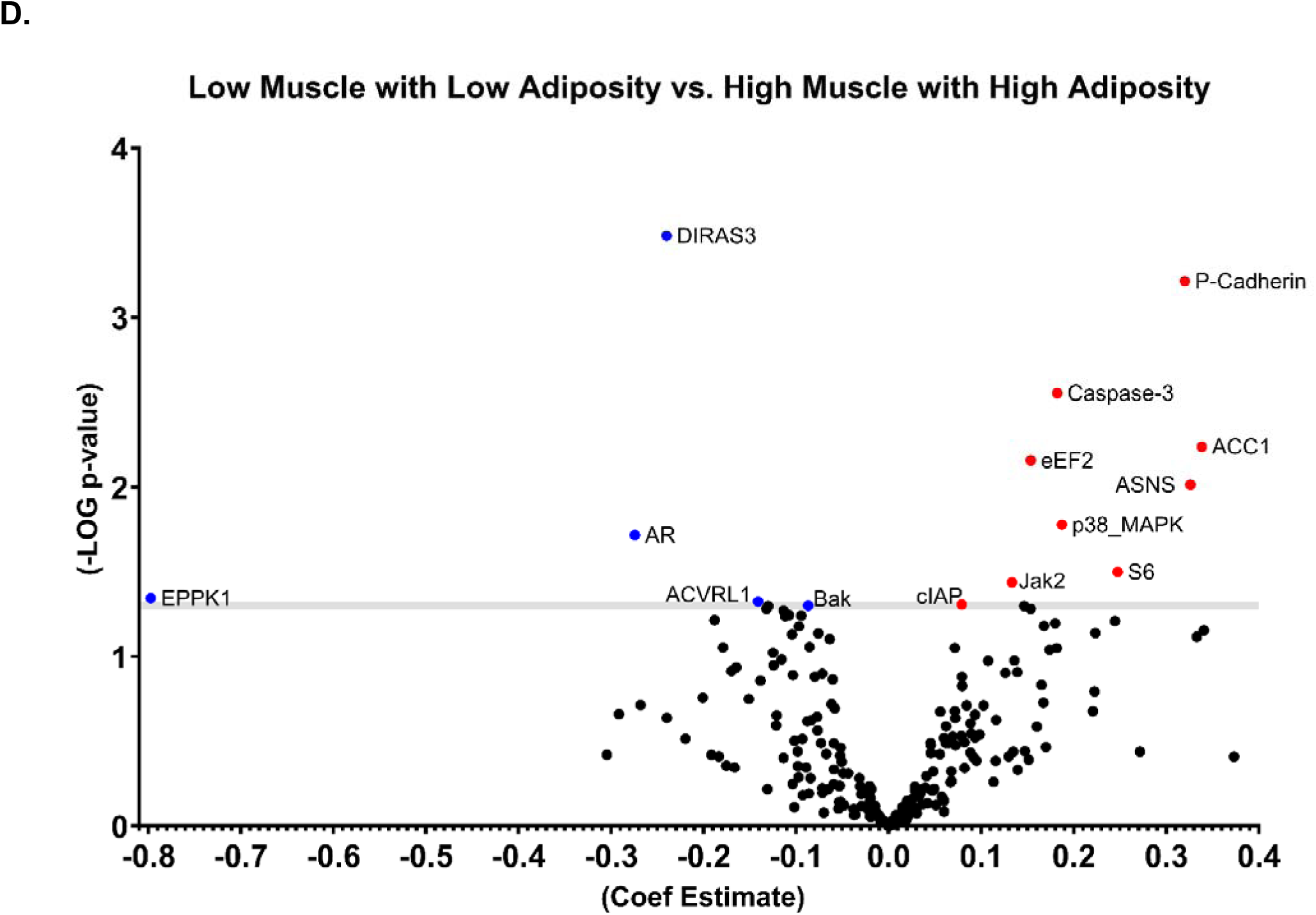
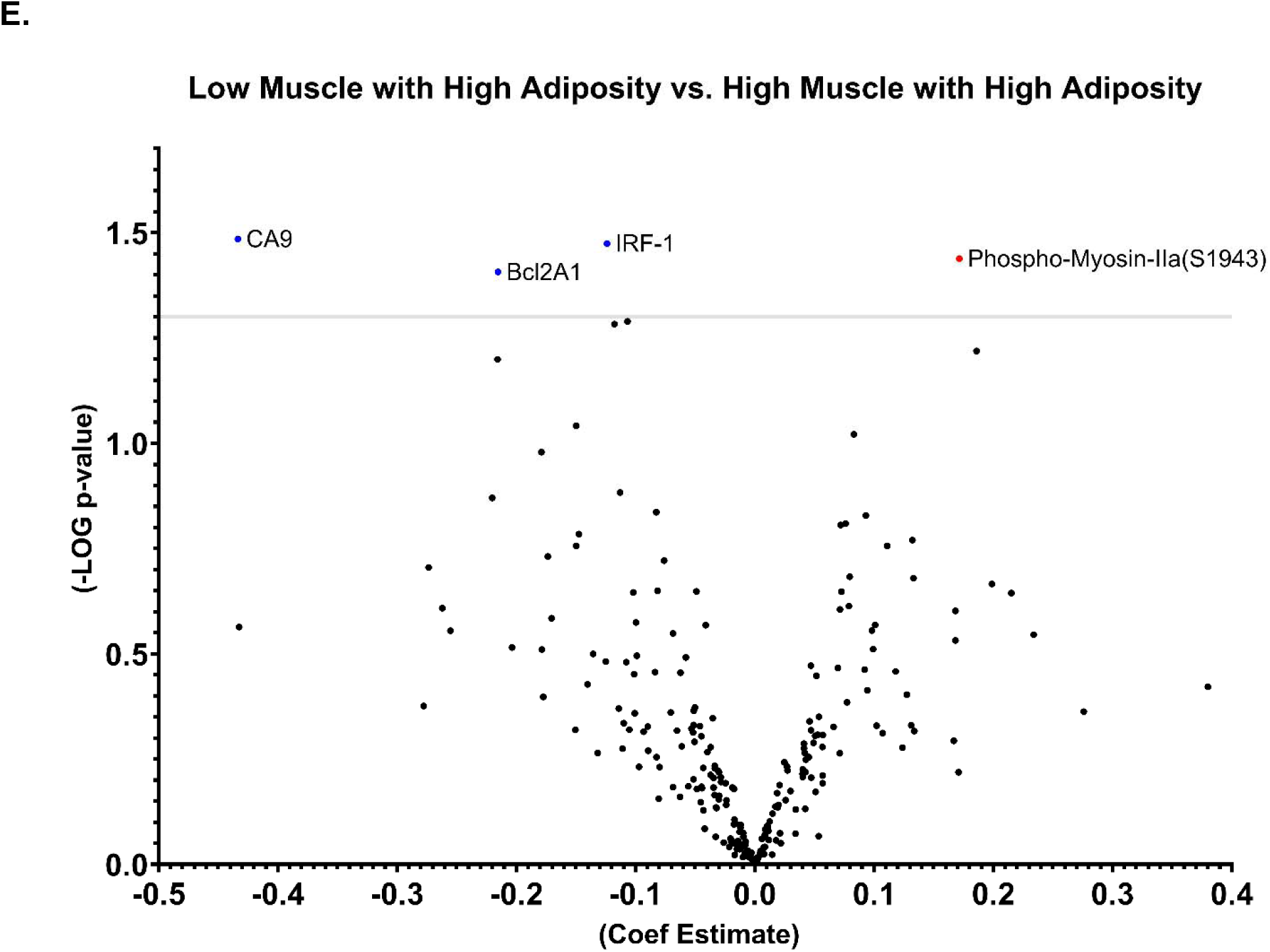
Volcano Plot of Differential Tumor Protein Expression Based on Total Skeletal Muscle, Total Adiposity Categories, and Body Composition Group.

High vs low TAT levels were associated with higher expression of DIRAS family GTPase 3 (DIRAS3) (log_2_ fold change, 0.13; P = 0.010), CD31 (log_2_ fold change, 0.1; P = 0.020), and cyclin D1 (log_2_ fold change, 0.1; P = 0.035), but lower expression of neurofibromin-2 (NF2) (log_2_ fold change, −0.1; P = 0.043), acetyl-CoA carboxylase 1 (ACC1) (log_2_ fold change, −0.2; P = 0.017), asparagine synthetase (ASNS) (log_2_ fold change, −0.2; P = 0.024), p38 mitogen-activated protein kinase (p38 MAPK) (log_2_ fold change, −0.2; P = 0.007), cellular inhibitor of apoptosis protein (cIAP) (log_2_ fold change, −0.1; P = 0.037), excision repair cross-complementation group 5 (ERCC5) (log_2_ fold change, −0.1; P = 0.028), phosphorylated acetyl-CoA carboxylase at serine 79 (phospho-ACC [S79]) (log_2_ fold change, −0.2; P = 0.037) and phosphatidylinositol 3,4,5-trisphosphate-dependent Rac exchanger 1 (PREX1) (log_2_ fold change, −0.1; P = 0.048) (**Figure 2B**).

**Table 3** presents the adjusted regression analysis findings, including proteins that exhibited at least one significant association with body composition (P < 0.05). **Supplemental File 1** contains the adjusted regression analysis for all 233 proteins. The high muscle with low adiposity type (vs. the high muscle with high adiposity type) was associated with higher expression of NF2 (log_2_ fold change, 0.2; P = 0.022) and lower expression of liver kinase B1 (LKB1) (log_2_ fold change, −0.1; P = 0.037) (**Figure 2C**).

**Table 3.** Differential Protein Expression by Body Composition and Tissue Quantity Category.

| Protein | H.M. with L.A. vs. H.M. with H.A. |  |  | L.M. with L.A. vs. H.M. with H.A. |  |  | L.M. with H.A. vs. H.M. with H.A. |  |  | Total Adiposity (High vs Low) |  |  | Skeletal Muscle (High vs Low) |  |  |
| --- | --- | --- | --- | --- | --- | --- | --- | --- | --- | --- | --- | --- | --- | --- | --- |
|  | Estimate | P-value | FDR | Estimate | P-value | FDR | Estimate | P-value | FDR | Estimate | P-value | FDR | Estimate | P-value | FDR |
| Phospho-ACC(S79) | 0.143 | 0.195 | 0.99 | 0.174 | 0.091 | 0.56 | -0.015 | 0.883 | 0.98 | 0.167 | 0.037* | 0.89 | 0.004 | 0.967 | 0.99 |
| ACC1 | 0.16 | 0.22 | 0.99 | 0.339 | 0.006* | 0.32 | 0.051 | 0.672 | 0.98 | 0.227 | 0.017* | 0.89 | 0.102 | 0.303 | 0.99 |
| AR | -0.042 | 0.738 | 0.99 | -0.274 | 0.019* | 0.55 | -0.216 | 0.063 | 0.98 | -0.051 | 0.576 | 0.96 | -0.222 | 0.02* | 0.99 |
| Bak | -0.014 | 0.769 | 0.99 | -0.087 | 0.05* | 0.55 | 0.025 | 0.572 | 0.98 | -0.065 | 0.061 | 0.89 | -0.014 | 0.690 | 0.99 |
| CD31 | -0.118 | 0.106 | 0.99 | -0.132 | 0.052 | 0.55 | -0.002 | 0.97 | 0.98 | -0.124 | 0.019* | 0.89 | -0.007 | 0.898 | 0.99 |
| clAP | 0.032 | 0.46 | 0.99 | 0.079 | 0.049* | 0.55 | -0.017 | 0.662 | 0.98 | 0.065 | 0.037* | 0.89 | 0.009 | 0.794 | 0.99 |
| Cyclin_D1 | -0.05 | 0.356 | 0.99 | -0.086 | 0.088 | 0.56 | 0.027 | 0.586 | 0.98 | -0.082 | 0.035* | 0.89 | 0.002 | 0.966 | 0.99 |
| eEF2 | -0.005 | 0.932 | 0.99 | 0.154 | 0.007* | 0.32 | 0.019 | 0.732 | 0.98 | 0.068 | 0.126 | 0.90 | 0.075 | 0.106 | 0.99 |
| LKB1 | -0.085 | 0.037* | 0.99 | -0.021 | 0.581 | 0.93 | -0.041 | 0.27 | 0.98 | -0.03 | 0.3147 | 0.90 | 0.001 | 0.974 | 0.99 |
| NF2 | 0.225 | 0.022* | 0.99 | 0.168 | 0.066 | 0.55 | 0.098 | 0.278 | 0.98 | 0.144 | 0.043* | 0.89 | 0.036 | 0.632 | 0.99 |
| P Cadherin | 0.047 | 0.634 | 0.99 | 0.32 | 0.001* | 0.071** | 0.101 | 0.27 | 0.98 | 0.137 | 0.058 | 0.89 | 0.17 | 0.025* | 0.99 |
| p38_MAPK | 0.143 | 0.087 | 0.99 | 0.187 | 0.017* | 0.55 | 0.002 | 0.981 | 0.98 | 0.165 | 0.007* | 0.89 | 0.019 | 0.765 | 0.99 |
| S6 | -0.078 | 0.528 | 0.99 | 0.247 | 0.032* | 0.55 | -0.015 | 0.893 | 0.98 | 0.1 | 0.266 | 0.90 | 0.121 | 0.201 | 0.99 |
| ASNS | 0.013 | 0.925 | 0.99 | 0.326 | 0.01* | 0.37 | -0.09 | 0.47 | 0.98 | 0.224 | 0.024* | 0.89 | 0.072 | 0.486 | 0.99 |
| Phospho-Myosin-IIa(S1943) | 0.004 | 0.961 | 0.99 | 0.126 | 0.125 | 0.60 | 0.172 | 0.036* | 0.98 | -0.022 | 0.733 | 0.97 | 0.152 | 0.024* | 0.99 |
| EPPK1 | -0.281 | 0.508 | 0.99 | -0.797 | 0.045* | 0.55 | -0.433 | 0.273 | 0.98 | -0.325 | 0.292 | 0.90 | -0.466 | 0.150 | 0.99 |
| ACVRL1 | -0.053 | 0.483 | 0.99 | -0.141 | 0.047* | 0.55 | -0.051 | 0.467 | 0.98 | -0.072 | 0.189 | 0.90 | -0.066 | 0.254 | 0.99 |
| DIRAS3 | -0.073 | 0.298 | 0.99 | -0.24 | <.001* | 0.071** | -0.051 | 0.431 | 0.98 | -0.134 | 0.01* | 0.89 | -0.098 | 0.071 | 0.99 |
| PREX1 | 0.128 | 0.11 | 0.99 | 0.147 | 0.05* | 0.55 | 0.043 | 0.564 | 0.98 | 0.115 | 0.048* | 0.89 | 0.033 | 0.587 | 0.99 |
| Bcl2A1 | -0.042 | 0.71 | 0.99 | -0.165 | 0.115 | 0.60 | -0.215 | 0.039 | 0.98 | 0.007 | 0.935 | 0.98 | -0.178 | 0.038* | 0.99 |
| Caspase_3 | -0.003 | 0.958 | 0.99 | 0.182 | 0.003* | 0.22 | 0.073 | 0.225 | 0.98 | 0.056 | 0.237 | 0.90 | 0.118 | 0.018* | 0.99 |
| ERCC5 | -0.003 | 0.964 | 0.99 | 0.079 | 0.149 | 0.63 | -0.107 | 0.051 | 0.98 | 0.096 | 0.028* | 0.89 | -0.031 | 0.495 | 0.99 |
| IRF-1 | 0.042 | 0.506 | 0.99 | -0.027 | 0.641 | 0.95 | -0.124 | 0.034 | 0.98 | 0.07 | 0.121 | 0.90 | -0.102 | 0.034* | 0.99 |
| Jak2 | 0.058 | 0.392 | 0.99 | 0.133 | 0.036* | 0.55 | 0.08 | 0.207 | 0.98 | 0.056 | 0.258 | 0.90 | 0.078 | 0.133 | 0.99 |
| CA9 | 0.168 | 0.441 | 0.99 | -0.054 | 0.789 | 0.97 | -0.434 | 0.033 | 0.98 | 0.2782 | 0.079 | 0.90 | -0.3487 | 0.037* | 0.99 |

Only associations with P<0.05 in the adjusted model. Models adjusted for stage, sex, race, ethnicity, and age
\* p-value < 0.05; \*\* FDR < 0.2; Abbreviations: FDR, false discovery rate; H.M., High Muscle; H.A., High Adiposity; L.M., Low Muscle; L.A., Low Adiposity.

The low muscle with low adiposity type (vs. the high muscle with high adiposity type) was associated with increased expression of P-cadherin (log_2_ fold change, 0.3; P = 0.001), caspase-3 (log_2_ fold change, 0.2; P = 0.003), ACC1 (log_2_ fold change, 0.3; P = 0.006), eukaryotic translation elongation factor 2 (eEF2) (log_2_ fold change, 0.2; P = 0.007), ASNS (log_2_ fold change, 0.3; P = 0.009), p38 MAPK (log_2_ fold change, 0.2; P = 0.017), ribosomal protein S6 (S6) (log_2_ fold change, 0.2; P = 0.032), Janus kinase 2 (Jak2) (log_2_ fold change, 0.1; P = 0.036), and cIAP (log_2_ fold change, 0.1; P = 0.049), and with decreased expression of DIRAS3 (log_2_ fold change, −0.2; P = 0.0003), androgen receptor (log_2_ fold change, −0.3; P = 0.019), epiplakin (EPPK1) (log_2_ fold change, −0.8; P = 0.045), activin A receptor type II-like kinase 1 (ACVRL1) (log_2_ fold change, −0.1; P = 0.047), and Bcl-2 antagonist killer 1 (Bak) (log_2_ fold change, −0.1; P = 0.0499) (**Figure 2D**).

The low muscle with high adiposity type (vs. the high muscle with high adiposity type) was associated with higher expression of phospho-myosin IIa (S1943) (log_2_ fold change, 0.2; P = 0.036) and with lower expression of CA9 (log_2_ fold change, −0.4; P = 0.033), IRF-1 (log_2_ fold change, −0.1; P = 0.034), and BCL2A1 (log_2_ fold change, −0.2; P = 0.038) (**Figure 2E**). All FDRs exceeded 0.2, except for P-cadherin and DIRAS3 in the low muscle with low adiposity type (vs. the high muscle with high adiposity type).

### Tumor protein expression and survival

Only PREX1 significantly interacted with body composition to predict survival (joint interaction p-value = 0.011). The cutoff value categorizing patients into high and low PREX1 groups and the corresponding survival curve are presented in **Supplemental Figure S4.** Tumor PREX1 high (vs. low) expression was associated with a higher mortality risk (HR, 2.12 [95% CI, 1.17–3.83]) (**Table 4**). Among the body composition strata, PREX1 high (vs. low) expression was associated with increased mortality risk among the patients having high muscle with high adiposity (HR, 2.99 [95% CI, 1.19– 7.51]) as well as a 15-fold higher mortality risk (HR, 15.77 [95% CI, 3.08–80.78) among patients having the low muscle with high adiposity type.

**Table 4:** PREX1 Protein Expression and Its Interaction with Body Composition on Survival

| **Tumor protein** | **HR** | **95% Confidence Interval** |  |
| --- | --- | --- | --- |
| PREX1 (high vs. low expression) | 2.12 | 1.17 | 3.83 |
| **Body Composition type** |  |  |  |
| High Muscle with Low Adiposity | 4.53 | 0.73 | 28.00 |
| High Muscle with High Adiposity* | 2.99 | 1.19 | 7.51 |
| Low Muscle with Low Adiposity | 0.79 | 0.33 | 1.90 |
| Low Muscle with High Adiposity* | 15.77 | 3.08 | 80.78 |
| Interaction Joint test P value, 0.011 |  |  |  |
| * Significant interaction $P < 0.05$.<br>Models were adjusted for age, sex, race, ethnicity, and cancer stage.<br>Abbreviations: HR, hazard ratio, PREX1, phosphatidylinositol 3,4,5-trisphosphate-dependent Rac exchanger 1 |  |  |  |

## Discussion

This study extends the findings of previous studies that evaluated the associations between body composition features and survival among patients with ccRCC (14–18) to include how body composition and its components interact with tumor protein expression in survival among patients with ccRCC. After adjusting for confounding factors, we found that the total area of adiposity (high vs. low TAT) as well as the body composition types of high muscle with low adiposity (vs. high muscle with high adiposity) and low muscle with low adiposity (vs. high muscle with high adiposity) were significantly associated with overall survival. We also found that several tumor proteins involved in carcinogenesis, functioning as oncoproteins or tumor suppressors in key cancer signaling pathways, were significantly upregulated or downregulated in high (vs. low) TSM, high (vs. low) TAT, and all body composition groups compared with the high muscle with high adiposity group. Body composition also modified the association between PREX1 expression levels and overall survival.

Patients with ccRCC displaying low adiposity (TAT low) exhibited overall mortality risk that was increased by more than twice that of patients with high adiposity (TAT high). This finding is consistent with the results of several previous studies and aligns with the "obesity paradox" observed for various obesity-associated malignant tumors, including renal cancers, in which higher adiposity is associated with better survival outcomes despite being a risk factor (11–13,29). The apparent obesity paradox may be explained by several factors, including that adipose tissue acts as an energy reserve, enabling patients with high adiposity to better withstand the physiological stress of cancer and the pharmacologic stress of cancer treatments. In addition, although we adjusted for cancer stage in our regression models, most patients with early-stage cancer were categorized not only in the high adiposity category but also in the high muscle category.

For the association between the differential expression of protein markers and adipose tissue, low TAT (vs. high TAT) was associated with a distinct molecular profile characterized by decreased expression of DIRAS3, CD31, and cyclin D1 alongside increased expression of several proteins, including NF2, ACC1, ASNS, p38 MAPK, cIAP, ERCC5, phospho-ACC (S79), and PREX1. This finding suggests that adipose tissue modulates tumor-suppressive and oncogenic mechanisms. Notably, DIRAS3, a tumor suppressor downregulated in patients with ccRCC having low TAT, binds to and disrupts RAS activation, thus inhibiting the RAS/MAPK pathway and limiting tumor growth. DIRAS3 also slows tumor progression and induces autophagy, which can support tumor dormancy and reduce tumor proliferative capacity (30). Thus, lower DIRAS3 expression in patients with low TAT is consistent with their poorer survival.

When we controlled for potential confounding factors, we observed no significant association between total muscle and overall survival. This finding contrasts with those of previous studies reporting that among patients with ccRCC, those with higher muscle mass exhibit significantly longer overall survival than those with lower muscle mass (16–18). Additionally, sarcopenia, characterized by a marked reduction in lean mass and weakened muscle strength, has been associated with increased mortality among patients with ccRCC (31,32). The difference between our findings and those of previous studies may involve the body composition classification methods employed. Our method resulted in twice as many patients categorized in the high TSM group as in the low TSM group, and most of these patients had early-stage cancer. However, while total muscle mass alone was not associated with overall survival, the combination of low muscle mass with low adiposity was associated with lower overall survival compared with the high muscle with high adiposity group, after adjusting for patient and tumor characteristics. This low muscle mass with low adiposity body composition profile likely reflects poor nutritional status at baseline, which has been linked to diminished benefits from medical treatments and lower overall survival rates for various types of cancer (33). Another group associated with high mortality in our study was the high muscle with low adiposity group. This finding underscores the importance of adiposity as a prognostic factor among patients with ccRCC, and it is consistent with our observation that high (vs. low) total adiposity is associated with better overall survival.

Tumors derived from patients with ccRCC having low muscle mass and/or low adiposity showed distinct and overlapping proteomic alterations associated with host body composition phenotypes. In patients with low muscle mass, tumors showed downregulation of androgen receptor, CA9, IRF1, and BCL2A1 proteins, alongside upregulation of P-cadherin, caspase-3, and phospho-myosin-IIA (S1943) proteins, indicating suppression of hormone and immune signaling with concomitant activation of apoptotic and cytoskeletal remodeling pathways. In contrast, tumors derived from patients with low adiposity exhibited increased expression of proteins related to metabolic regulation and cellular stress, including NF2, ACC1, phospho-ACC (S79), ASNS, p38 MAPK, ERCC5, cIAP, and PREX1, together with reduced expression of DIRAS3, CD31, and cyclin D1, suggesting dysregulated lipid metabolism, enhanced stress kinase signaling, impaired angiogenesis, and altered cell cycle control.

Interestingly, tumors derived from patients presenting with both low muscle mass and low adiposity uniquely displayed a common proteomic signature characterized by upregulation of JAK2, ribosomal S6, and eEF2, reflecting sustained activation of inflammatory and mammalian target of rapamycin signaling, along with downregulation of EPPK1, ACVRL1, and pro-apoptotic BAK, suggesting disruption of epithelial structure, angiogenic signaling, and apoptotic regulation. In particular, DIRAS3 and P-cadherin, proteins that were significantly associated with the low muscle with low adiposity type after FDR correction, may be key proteins linking body composition and survival. In our study, P-cadherin expression was upregulated in the low muscle with low adiposity type. This upregulation is consistent with the survival data in the TCGA-KIRC cohort showing that high (vs. low) P-cadherin expression was associated with increased mortality risk (Log-Rank P, 2.468e-7) (34). Similarly, DIRAS3 expression was downregulated in the low muscle with low adiposity type in our study, and in the TCGA-KIRC cohort, low (vs. high) DIRAS3 expression was associated with increased mortality risk, although the association was not significant (Log-Rank P, 0.153) (34).

These shared molecular alterations likely reflect systemic features of cachexia (35), such as chronic inflammation, energy imbalance, and host metabolic stress, which broadly impact tumor biology regardless of specific tissue depletion. In contrast, distinct proteomic profiles between low muscle and low adiposity phenotypes may result from the differential roles of skeletal muscle and adipose tissue in host–tumor crosstalk. Loss of muscle mass appears to primarily affect hormone, cytokine, and anabolic signaling pathways, whereas fat depletion influences nutrient sensing, lipid metabolism, and angiogenic factors. Together, these distinct and overlapping alterations underscore how body composition phenotypes shape the tumor proteome through both shared systemic stressors and tissue-specific host factors.

Our analysis revealed that body composition modified the association between PREX1 and survival. High expression of PREX1 in patients having low muscle with high adiposity was associated with a 15.8-fold increase in mortality risk. This finding is in agreement with previous studies, with high PREX1 expression consistently associated with poor prognosis across various cancers (36–38) and with preclinical models demonstrating that PREX1 knockdown hinders tumor growth and metastasis (36,37).

Located downstream of receptor tyrosine kinases and G protein-coupled receptors (39,40), PREX1 activates Rho GTPases, primarily Rac1, orchestrating cellular functions such as cytoskeletal remodeling and promoting cell migration and proliferation (40,41). We found that PREX1 was upregulated in tumors of patients with higher adiposity, in concordance with a recent study suggesting that PREX1 may promote glucose intolerance, as PREX1 knockout mice fed a high-fat diet were protected from developing diabetes (42). Overall, our findings speak to the prooncogenic and metabolic roles of PREX1. In patients with high adiposity, obesity-induced inflammation and metabolic deregulation may have enhanced the oncogenic effects of PREX1, leading to more aggressive tumor behavior, increased metastatic potential, and poorer survival.

High PREX1 expression was associated with even poorer survival when high adiposity coexisted with low muscle, typical of sarcopenic obesity (43,44). A low muscle with high adiposity phenotype along with high PREX1 expression may create a particularly vulnerable state that independently predicts poor cancer prognosis.

A major strength of this study is that our evaluation of the association between CT-assessed muscularity and adiposity in relation to tumor proteomics among patients with ccRCC highlighted distinct interactions of muscle vs. adipose tissue with tumor biology. However, this study has several limitations. The retrospective design may have introduced selection bias. Additionally, the analysis was cross-sectional using a single CT scan image obtained close to diagnosis; hence, establishing causal relationships between body composition and tumor proteomics was not possible. Systematic errors may have arisen from variability in CT scan images obtained from different centers. This variability could not be addressed in the analysis due to a lack of this information. In addition, the TCGA-KIRC cohort lacked weight and height data, which would have been valuable in model adjustment or comparison with the body composition variables.

## Conclusion

The body composition type of low muscle mass combined with low adiposity was associated with poor survival among patients with ccRCC. Low Muscle with low adiposity body composition type was associated with distinct tumor proteomic patterns, including increased expression of proteins related to cell proliferation and migration, and decreased expression of tumor suppressor proteins. The modification of body composition type on tumor PREX1 expression and its association with survival highlights the critical interactions between host body composition and tumor molecular characteristics, supporting the potential utility of integrating both types of factors in prognostic assessment and risk stratification of ccRCC.

## Data Availability

The CT images analyzed in this study were obtained from the Cancer Imaging Archive at TCIA. Clinical and protein data are available in the National Cancer Institute Genomic Data Commons and at MD Anderson Cancer Center The Cancer Proteome Atlas, respectively. Data generated by the authors is available upon request.

