## Supplemental Tables and Figures for "Association of Body Composition With Tumor Proteomics and Survival in Patients With Clear Cell Renal Cell Carcinoma"

**Supplemental Table S1****: Tissue radiodensity cutoff points in Hounsfield units for computed tomography image annotation**

| **Tissue** | **Tagged tissue locations** | **Lower limit of Hounsfield unit** | **Upper limit of Hounsfield unit** |
| --- | --- | --- | --- |
| Subcutaneous adipose tissue | Subcutaneous fat | -190 | -30 |
| Intermuscular Adipose tissue | Intermuscular fat | -190 | -30 |
| Visceral adipose tissue | Fat around organs | -150 | -50 |
| Total skeletal muscle | Skeletal muscle | -29 | 200 |
| Bone | Bone | 200 | 2001 |

**Supplemental Table S2: Hazard of mortality for covariates**

| **Variable** | **HR (95% CI)** | **P-Value** |
| --- | --- | --- |
| Sex (Reference: Female) | 2.28 (1.16-4.45) | 0.016^1^ |
| Age: Step Size: 10 years | 1.32 (1.03-1.70) | 0.027^2^ |
| Race (Reference: White) |  | 0.714^1^ |
| Asian | 1.23 (0.16-9.55) | 0.842^2^ |
| Black or African American | 0.44 (0.06-3.34) | 0.424^2^ |
| Ethnicity (Reference: Hispanic or Latino) |  | <.001^1^ |
| Not Hispanic or Latino | 5.20 (0.69-39.31) | 0.110^2^ |
| Not Reported | 16.31 (2.04-130.17) | 0.008^2^ |
| AJCC Pathologic Stage (Reference: Stage I) |  | <.001^1^ |
| II | 0.93 (0.26-3.35) | 0.912^2^ |
| III | 3.77 (1.85-7.71) | <.001^2^ |
| IV | 8.46 (4.12-17.37) | <.001^2^ |
| ^1^Type 3 Wald p-value; ^2^Covariate Wald p-value.  All models were adjusted for all other covariates and body composition groups.  Abbreviations: AJCC, American Joint Committee on Cancer; CI, confidence interval; HR, hazard ratio. | | |

**Supplemental Figure S1: Flowchart of patient selection from the TCGA-KIRC cohort**

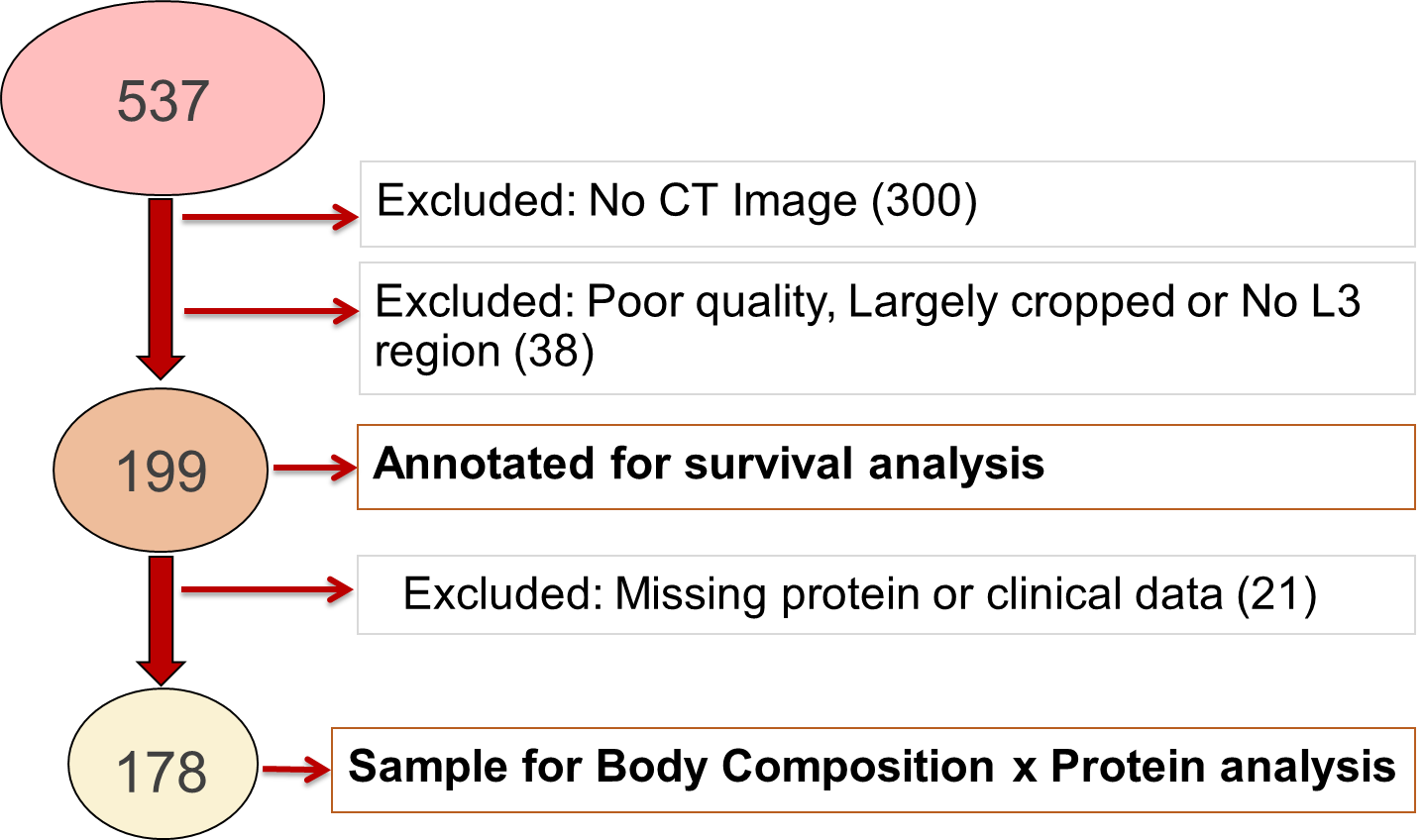

**Abbreviations: CT, computer tomography; L3, third lumbar vertebra**

**Supplemental Figure S2: Total muscle area cutoffs**

A. **FEMALE**

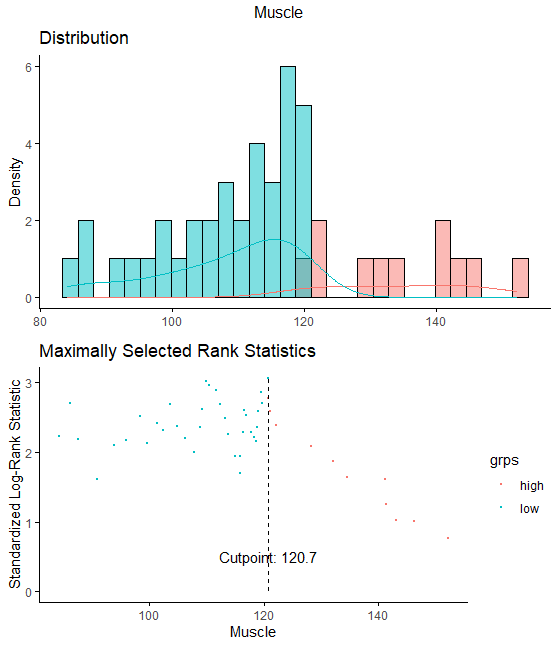

B. **MALE**

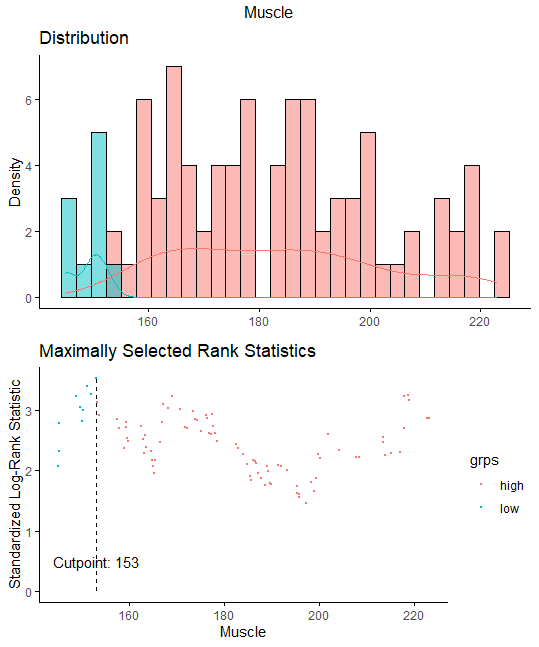

**Supplemental Figure S3: Total adipose tissue cutoffs**

A. **FEMALE**

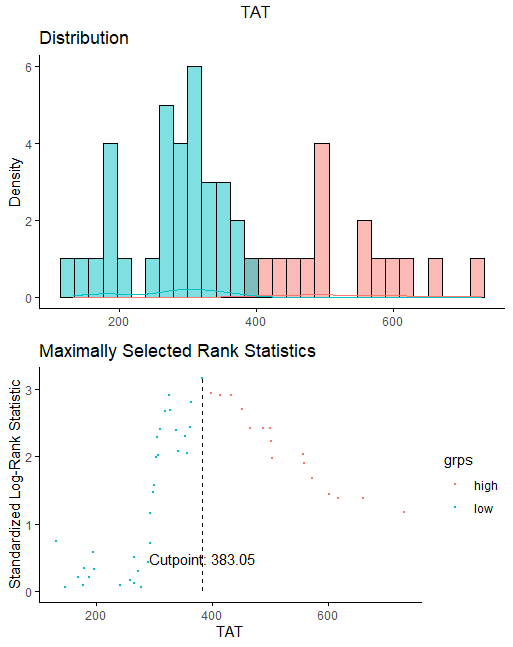

B. **MALE**

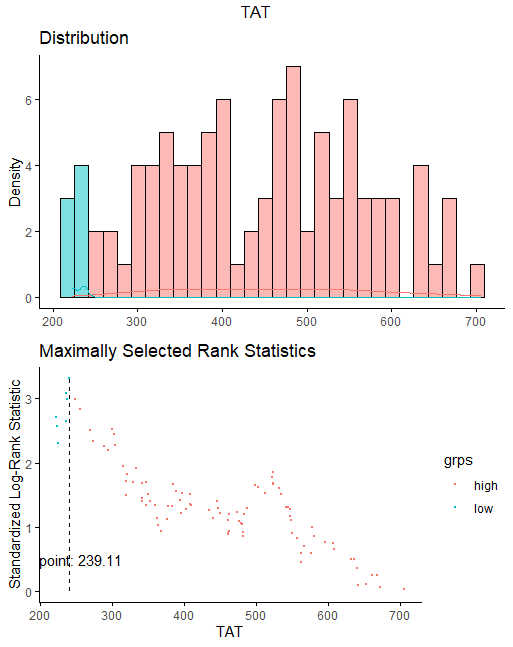

**Supplemental Figure S4: PREX1 distribution and corresponding Kaplan-Meier survival curve**

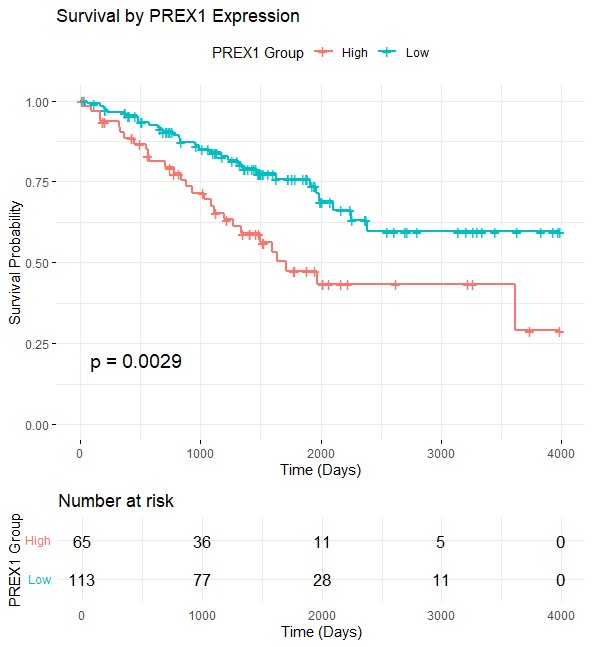

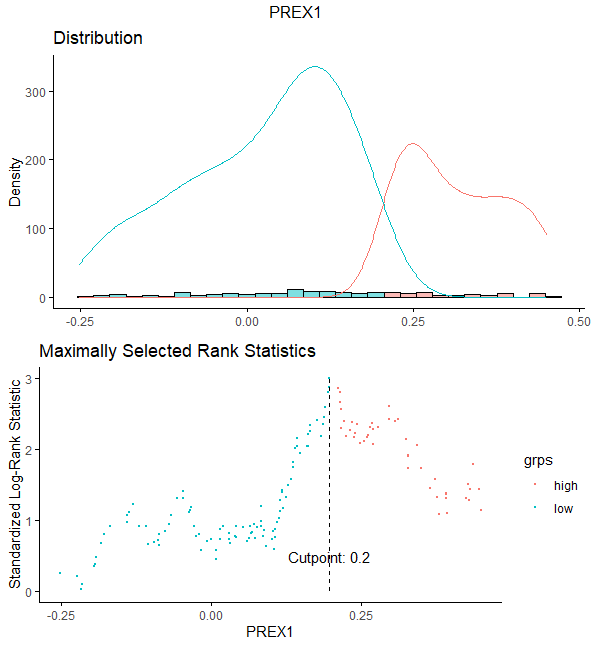
